# A novel ultrasensitive assay for plasma p-tau217: performance in individuals with subjective cognitive decline and early Alzheimer’s disease

**DOI:** 10.1101/2023.09.26.23296134

**Authors:** Fernando Gonzalez-Ortiz, Pamela C L Ferreira, Armand Gonzalez, Laia Montoliu-Gaya, Paula Ortiz-Romero, Przemyslaw R. Kac, Michael Turton, Hlin Kvartsberg, Nicholas J. Ashton, Henrik Zetterberg, Peter Harrison, Bruna Bellaver, Guilherme Povala, Victor L. Villemagne, Tharick A. Pascoal, Mary Ganguli, Anne D. Cohen, Carolina Miguillon, Jose Contador, Marc Suarez-Calvet, Thomas K. Karikari, Kaj Blennow

## Abstract

**INTRODUCTION:** Detection of Alzheimer’s disease (AD) pathophysiology among cognitively unimpaired individuals and those experiencing subjective cognitive decline (SCD) remains challenging. Plasma p-tau217 is one of the most promising of the emerging biomarkers for AD. However, accessible methods are limited.

**METHODS:** We employed a novel p-tau217 immunoassay (UGOT p-tau217) in four independent cohorts (n=308) including a cerebrospinal fluid (CSF) biomarker-classified cohort (Discovery), two cohorts consisting mostly of cognitively unimpaired participants (MYHAT and Pittsburgh), and a population-based cohort of individuals with SCD (β-AARC).

**RESULTS:** UGOT p-tau217 showed high accuracy (AUC= 0.80-0.91) identifying Aβ pathology, determined either by Aβ positron emission tomography or CSF Aβ42/40 ratio. In individuals experiencing SCD, UGOT p-tau217 showed high accuracy identifying those with a positive CSF Aβ42/40 ratio (AUC= 0.91).

**DISCUSSION:** UGOT p-tau217 can be an easily accessible and efficient way to screen and monitor patients with suspected AD pathophysiology, even in the early stages of the continuum.

## 1. Background

Alzheimer’s disease (AD) poses a significant global health challenge with an increasing prevalence as the population ages^1–3^. Meanwhile, the evaluation of individuals in early stages of the AD continuum continues to be difficult partly due to heterogeneity in how clinical manifestations compare with biological evidence of disease^4, 5^. The implementation of peripheral biomarkers that can provide biochemical evidence of early AD biological changes could help clinicians to provide specialised diagnostic and/or therapeutic care^2, 3^. In the era of new anti-amyloid-beta (Aβ) immunotherapy drugs for AD, early intervention will be critical for maximising treatment efficacy, and robust biomarkers that become abnormal in cognitively unimpaired individuals as well as patients in the early symptomatic phases will be essential for first-in-line screening of patients eligible for treatment^5, 6^.

While neuroimaging techniques, especially positron emission tomography (PET), and cerebrospinal fluid (CSF) analyses offer valuable insights into AD pathology, they are expensive, invasive, involve radioactive ligands, and have very limited accessibility for routine clinical use^7, 8^. Therefore, the search for minimally invasive and readily accessible biomarkers for AD has gained substantial momentum^2, 9^. Several plasma biomarkers have been shown to associate well with Aβ pathophysiology – whether assessed using Aβ PET or CSF Aβ42/40 ratio^8, 10^ – as well as cognitive performance^11, 12^. These high-performing biomarkers include plasma Aβ42/40 measured with immunoprecipitation-mass spectrometry, glial fibrillary acidic protein (GFAP), and p-tau variants^13–16^. Among the plasma p-tau biomarkers, p-tau181, p-tau217 and p-tau231 have all shown high performances to detect Aβ pathophysiology^2, 3^. However, recent head-to-head studies suggest that p-tau217 has the largest between-diagnostic group fold increases and the strongest associations with both cross-sectional and longitudinal changes in Aβ pathophysiology^17^, explainable by a more pronounced longitudinal increase in symptomatic cases^18^. The diagnostic accuracy and sensitivity of plasma p-tau217 have been consistently demonstrated across different publications by various research groups, outperforming other blood-based biomarkers^15, 17, 19–22^. While further studies in other cohorts are needed, the current evidence suggests that p-tau217 is the most promising of the plasma p-tau biomarkers presently available.

Despite its outstanding diagnostic capabilities, available plasma p-tau217 assays are currently accessible only from a selected group of pharmaceutical/biotechnology companies; currently, no academic institution has developed a functional immunoassay for plasma p-tau217. To this end, only a handful of cohorts have been evaluated using plasma p-tau217 when compared with p-tau181 for which commercial assays exist^17^. This limited access has several drawbacks. For example, plasma p-tau217 has mostly been evaluated in cohorts with advanced characterization using concurrent CSF assays and neuroimaging methods. However, plasma p-tau217 assessment in cohorts that are more reflective of the “real world” population such as population-based studies as well as those with subjective cognitive decline who make up a substantial proportion of memory clinic patients are lacking.

Here, we sought to address these limitations by developing a plasma p-tau217 assay at the University of Gothenburg (UGOT), Sweden, that we refer to as UGOT p-tau217. We employed a newly generated sheep monoclonal antibody to develop a Simoa-based assay for plasma p-tau217 in blood. In this multicentre study, we present the technical validation of the UGOT p-tau217 assay and its clinical performance, focusing on individuals in early symptomatic stages of the disease, namely SCD or MCI stage^23, 24^.

## 2. Methods

### 2.1 Study cohorts, design, and outcome

#### Discovery cohort

The discovery cohort (n=40) included paired CSF and plasma samples from neurochemically defined AD dementia patients (n=20) and age-matched controls (n=20) from the Sahlgrenska University Hospital, Gothenburg, Sweden. The AD patients were selected based on their core CSF biomarker profile, according to reference values previously described (CSF Aβ42 <530 ng/L, p-tau181 >60 ng/L, and total-tau >350 ng/L)^13^, and had no evidence of other neurological conditions based on routine clinical and laboratory assessments. The control group consisted of selected patients without an AD profile by clinical evaluation and CSF biomarkers.

#### MYHAT cohort

The Monongahela-Youghiogheny Healthy Aging Team (MYHAT; n= 79) is a research cohort that focuses on a group from southwestern Pennsylvania, USA^25^. The participants were selected through random sampling from voter registration lists during two time periods: 2006-2008 and 2016-2019. To be eligible, participants had to be at least 65 years old, live in a specific town within the target area, not reside in long-term care facilities, have adequate hearing and vision for testing, and be able to make decisions. Participants were classified as cognitively unimpaired (CU) if they had a Clinical Dementia Rating (CDR) of 0, mildly cognitively impaired (MCI) if they had CDR of 0.5, and with dementia if they had a CDR of ≥1. The analysis in the MYHAT cohort focused on cognitively normal participants and those with early cognitive changes (CDR 0 and CDR 0.5 respectively). The study was approved by the University of Pittsburgh Institutional Review Board and all participants gave written informed consent.

#### Pittsburgh cohort

The Pittsburgh cohort (n = 93) consisted of volunteers who participated in one of the following research studies conducted at the University of Pittsburgh: The Heart Strategies Concentrating on Risk Evaluation (HEART Score) parent study^26^ and The Human Connectome Project (HCP)^27^, The Normal Aging study^11^ and the MsBrain study^28^. All participants provided written informed consent. Participants were classified as CU if they had a CDR of 0, MCI if they had a CDR of 0.5, and with dementia if they had a CDR of ≥1. The amyloid status in these patients was evaluated using Aβ PET. The study was approved by the University of Pittsburgh Institutional Review Board and all participants gave written informed consent.

#### β-AARC cohort

The Barcelonaβeta Brain Research Center’s Alzheimer’s At-Risk Cohort (β-AARC) is a longitudinal, prospective, and observational cohort study for the early identification of blood-based biomarkers in a population with SCD that have sought / are seeking medical advice. Participants of the β-AARC study are comprehensively characterized and several relevant variables are collected, including, among others, clinical and cognitive variables (including SCD characterisation), magnetic resonance imaging (MRI) and CSF and blood biomarkers. The inclusion criteria include: (1) Cognitively unimpaired persons with SCD, as well as MCI, that have sought / are seeking medical advice; (2) Participation (in-person at the institution or telephonically) of a relative to inform on the participant’s SCD and on the clinical interview; (3) Men and women between 55 and 80 years old; and (4) Good knowledge of either Spanish or Catalan and being literate. Exclusion Criteria include: (1) Presence of clinically relevant psychiatric disorder according to the DSM-V criteria; (2) Multiple sclerosis, epilepsy in treatment and with frequent seizures (> 1 / month) in the last year, Parkinson’s disease or other neurodegenerative disease; (3) Contraindication to undergo MRI; (4) Contraindication to lumbar puncture (LP); (5) Acquired brain injury; (6) Investigator’s criteria: Subjects that show any condition that, in the opinion of the investigator, could interfere in the proper execution of the study procedures and / or in their future permanence in the study. The β-AARC study was approved by the independent ethics committee ‘Parc de Salut Mar’, Barcelona, and registered as Clinicaltrials.gov (identifier: NCT04935372).

### 2.2 Development and validation of the UGOT p-tau217 assay

A novel sheep monoclonal antibody, which selectively binds to tau phosphorylated specifically at threonine-217, was generated, characterized and used as the capture antibody. A mouse monoclonal antibody (Tau12, GenScript Biotech) raised against the N-terminal region of tau was used for detection. *In vitro* phosphorylated recombinant full-length tau-441 (#TO8-50FN, SignalChem) was used as the assay calibrator. Blood samples and calibrators were diluted with the assay diluent (Tau 2.0; #101556, Quanterix). Assay validation focused on within- and between-run stability, dilution linearity, spike recovery and determination of the lowest limit of quantification. Analytical validation followed protocols described previously^29, 30^. Assay development work was done at the University of Gothenburg, Sweden. The resulting assay is hereby referred to as UGOT p-tau217 since it originates from the University of Gothenburg.

### 2.3 Measurement of p-tau217 using the UGOT assay in the clinical cohorts

UGOT p-tau217 was measured blinded (without knowledge on the clinical data) on Simoa HD-X using the above-described in-house assay at the Department of Psychiatry and Neurochemistry, University of Gothenburg, Mölndal, Sweden. Signal variations within and between analytical runs were assessed using two internal quality control samples analysed in duplicates at the beginning and the end of each run.

### 2.4 Measurement of other plasma biomarkers

All biomarkers were measured on the Simoa HD-X platform. Plasma p-tau181 was measured either with a commercial method from Quanterix Inc. (p-tau181 V2 Advantage Kit #103714) or according to the Karikari *et al* method ^13^ for all other cohorts. Plasma brain-derived tau (BD-tau) was measured according to Gonzalez-Ortiz et al.^31^, and plasma p-tau231 by the published method by Ashton et al^14^. Measurement of p-tau217 by mass spectrometry was performed following the previously described protocol by Montoliu-Gaya et al. ^32^.

### 2.5 Aβ PET

In the Pittsburgh cohort, [11C]PiB PET acquired at 50–70 minutes post-injection was used to quantify Aβ PET uptake. We used a previously published method to transform the Aβ PET SUVR to the Centiloid scale^33^. The Aβ PET positivity was defined using a previously published cutoff^6^ with a cut-off of Centiloids ≥12 used to detect early Aβ aggregation in CU individuals.

### 2.6 Statistical analyses

Statistical analyses were performed with Prism version 9.3.1 (GraphPad, San Diego, CA, USA) and the R programming language. The distributions of data sets were examined for normality using the Kolmogorov-Smirnov or Shapiro-Wilks tests. Data are shown as mean ± standard deviation or median and IQR if non-normal. Non-parametric tests were used for non-normally distributed data. Spearman correlation and the χ2 test were used for continuous and categorical variables respectively. Diagnostic performances were evaluated with Receiver Operating Curves and Area Under the Curve (AUC) assessments. Fold changes were examined by comparing biomarker values with the mean of the control group. Group differences were examined using the Mann-Whitney test (two categories) or the Kruskal-Wallis test with Dunn’s multiple comparison (three or more groups).

## 3. Results

### 3.1 Analytical validation

Direct ELISA assessments showed that the new p-tau217-directed sheep monoclonal antibody was indeed specific for phosphorylation at threonine-217 and did not react with synthetic peptides that were unphosphorylated or were phosphorylated only at the neighbouring threonine-212 or threonine-214 sites (Figure S1). The analytical validation results showed that the assay signal diluted linearly in proportion to the fold dilution, signal from exogenously added material was recoverable with high accuracy, and aliquots of identical samples measured on different days gave highly precise readings (Figure S2). The lower limit of quantification for the assay (LLOQ) was estimated to be 0.08 pg/mL. LLOQ was calculated by serially diluting the highest assay calibrator point (53.7 pg/mL) two-fold (and in duplicates) and setting the LLOQ as the calibrator point immediately preceding the first concentration where the coefficient of variation (CV) was 20% or above.

### 3.2 UGOT p-tau217 in the Discovery cohort

The Discovery cohort included 14 (70%) and 12 (60%) females in the AD and control groups respectively. Demographic characteristics can be found in the supplementary appendix (Table S1). CSF and plasma UGOT p-tau217 concentrations were both higher in Aβ+ AD dementia versus Aβ-controls (Figure 1). While the UGOT p-tau217 fold change in CSF was higher, in comparison, CSF p-tau181 showed a lower fold change and a larger overlap between groups but was still significantly higher in the AD group compared with controls (P=0.001). Plasma UGOT p-tau217 showed strong correlations with CSF UGOT p-tau217 (R=0.80, p<0.001), CSF p-tau181 (R=0.77, p<0.001) and CSF total-tau (R=0.69, p<0.001) and was negatively correlated with the Aβ42 in CSF (R= −0.74, p<0.001).

**Figure 1.**
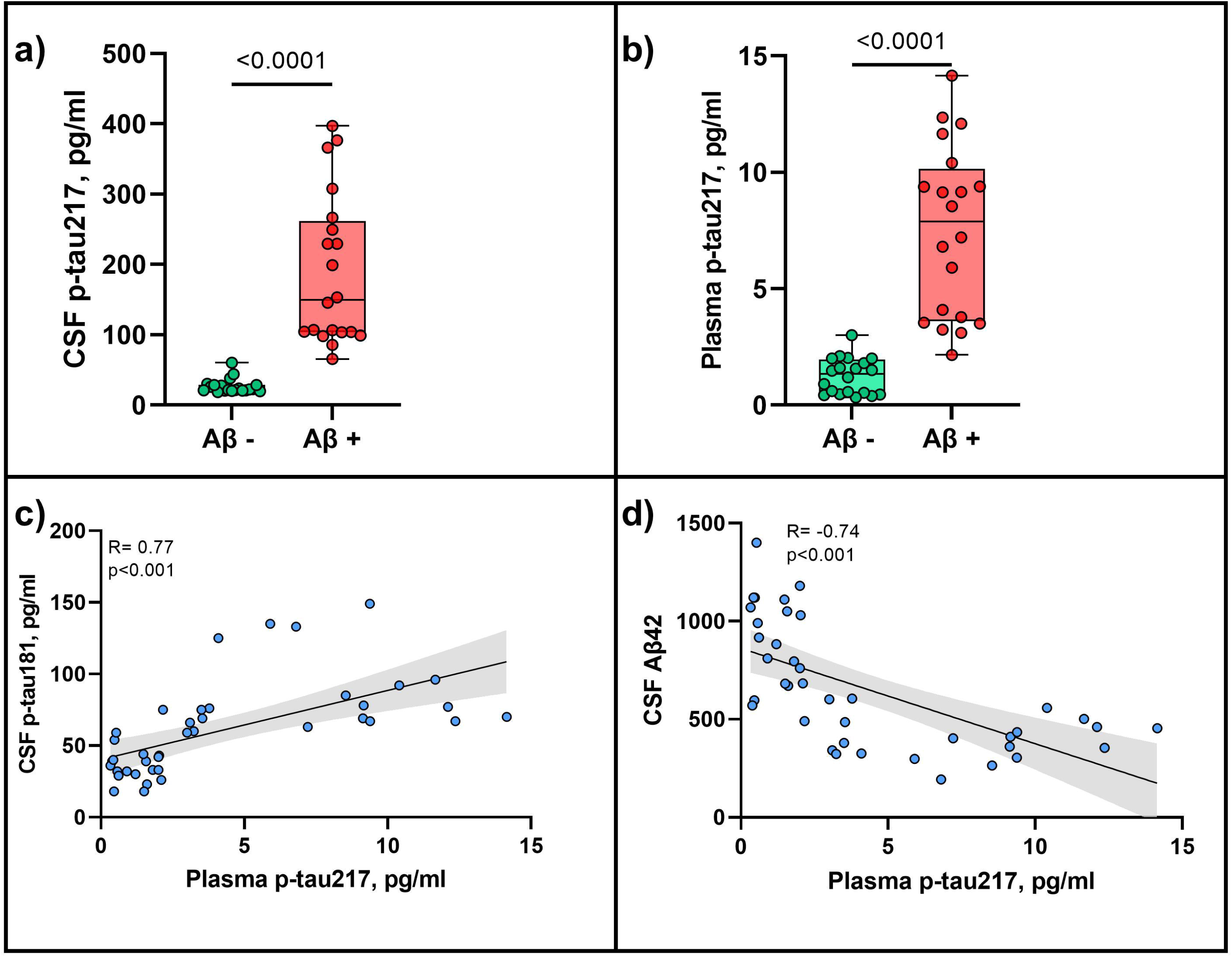
CSF and Plasma UGOT p-tau217 differentiate Aβ positive from Aβ negative individuals. *(1a-b)* UGOT p-tau 217 show high accuracy discriminating between biomarker-defined AD patients and age-matched controls (p<0.001) in CSF and plasma (p<0.001). (*1c-d)* Plasma UGOT p-tau217 showed a strong correlation with CSF p-tau181 (R=0.77, p<0.001) and negatively correlated with the amyloid 42 in CSF (R= −0.74, p<0.001).

### 3.3 UGOT p-tau217 in the population-based MYHAT cohort

The MYHAT cohort included 30 (75%) and 30 (76.9%) females in the CDR 0 and CDR 0.5 groups respectively. Demographic characteristics can be found in the supplementary appendix (Table S2). Plasma UGOT p-tau217 concentrations were higher in individuals with early cognitive decline (CDR=0.5) compared with those who were cognitively normal (CDR=0) with an AUC of 0.80, outperforming p-tau181 and GFAP (Figure 2a-b). The fold change for plasma UGOT p-tau217 was higher than for p-tau181 (Table S1), separating groups better than any of the other markers evaluated (plasma GFAP, p-tau181, BD-tau and NfL). Fold changes between CDR 0 and CDR 0.5 were greater for p-tau 217 (1.9) compared to p-tau181 (1.3). Moreover, UGOT p-tau217 levels showed a moderate correlation with plasma p-tau181 (R=0.51, p<0.001). Concerning correlations with neurodegeneration biomarkers, there was a stronger correlation with plasma BD-tau (R=0.68, p<0.001) than with plasma NfL (R=0.37, p<0.001; Fig. 2c-d).

**Figure 2.**
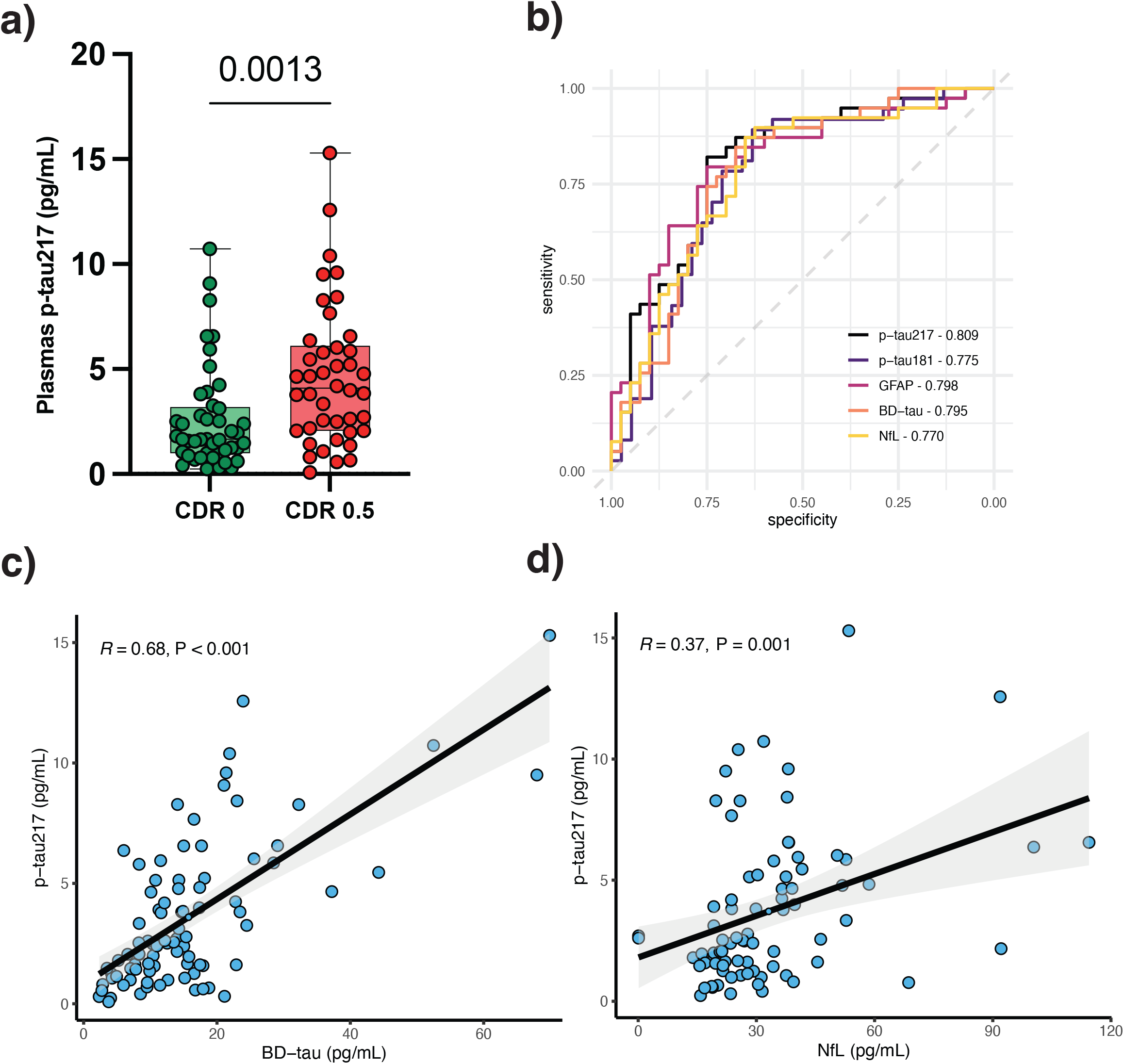
Plasma UGOT p-tau217 identifies individual with early cognitive changes. *(2a-b)* In the MYHAT cohort, plasma UGOT p-tau217 discriminated between participants with CDR 0 and CDR 0.5 (p=0.0013) with an AUC of 0.80. *(2c-d)* Plasma UGOT p-tau217 correlated strongly with plasma BD-tau (R=0.68, p<0.001) and weakly with plasma NfL (R=0.37, p<0.001).

### 3.4 Plasma UGOT p-tau217 associations with Aβ-PET uptake in the Pittsburgh cohort

The Pittsburgh cohort included 46 (63.9%) and 9 (42.9%) females in the Aβ PET Negative and Aβ PET Positive groups respectively. Demographic characteristics can be found in the supplementary appendix (Table S4). Plasma UGOT p-tau217 performed better than plasma p-tau231, p-tau181, GFAP and NfL to differentiate Aβ-PET positive and negative individuals with an AUC of 0.90 while the AUCs for the other biomarkers were between 0.79 – 0.81 (Fig. 3b). The fold change was largest for UGOT p-tau217 (2.8) compared with p-tau231 (1.07) and p-tau181 (1.15). Furthermore, plasma UGOT p-tau217 correlated strongly with Aβ-PET in Aβ positive individuals (R=0.57 p=0.007, Figure 3d).

**Figure 3.**
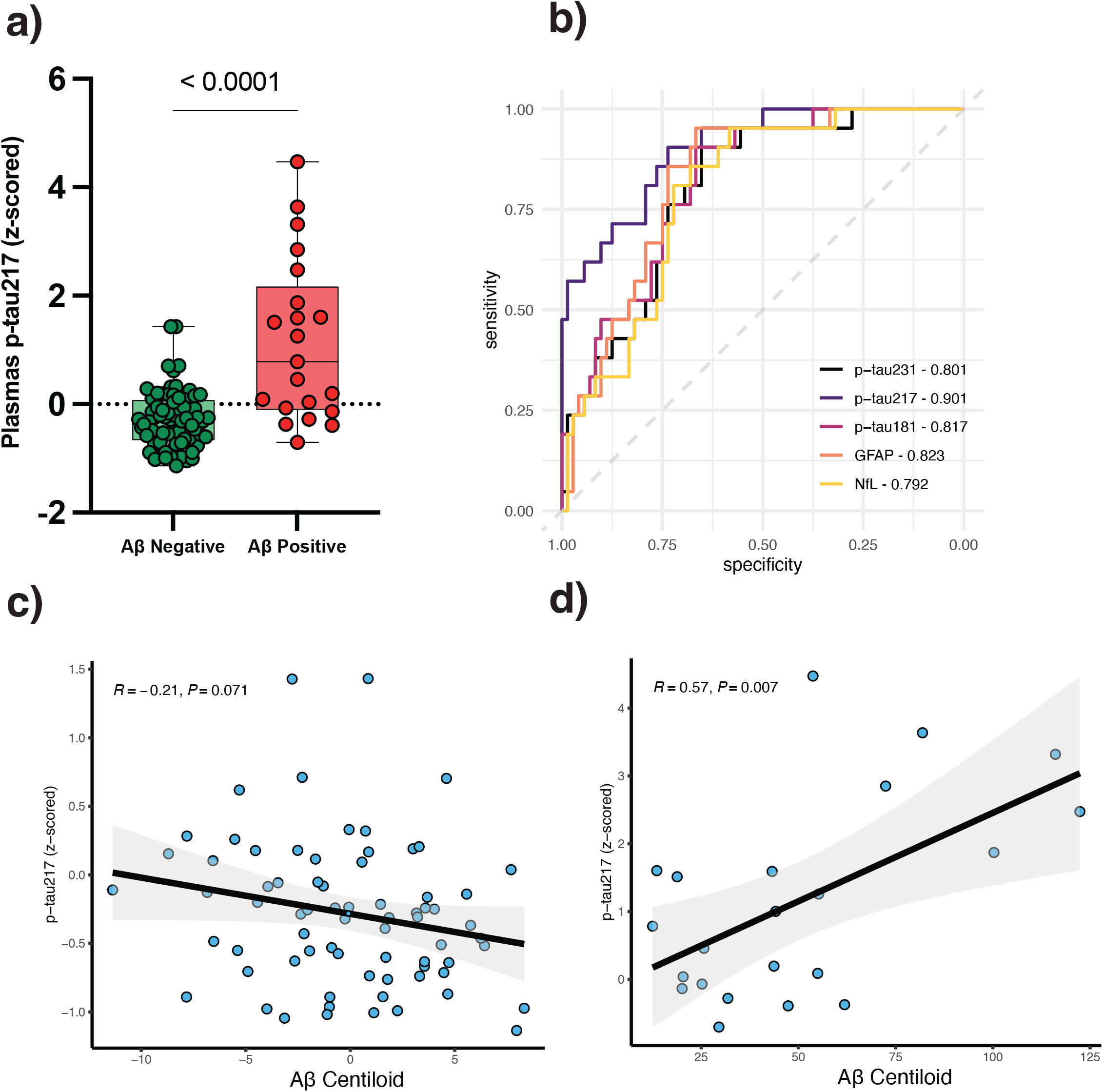
Plasma UGOT p-tau217 discriminate between Aβ PET positive and Aβ PET negative individuals in early AD. *(3a-b)* In the Pittsburgh cohort, plasma UGOT p-tau217 was able to differentiate Aβ+ from Aβ– individuals determined by Amyloid PET (p<0.001), showing an AUC of 0.90, outperforming p-tau181, p-tau231, GFAP and NfL. *(3c-d)* Plasma UGOT p-tau217 showed no correlation with Aβ PET in Aβ-individuals (R= −0.21, p=0.071) and strong correlation in Aβ+ individuals (R=0.57, p=0.007).

### 3.5 Plasma p-tau217 performance in the β-AARC cohort (early symptomatic individuals, cognitively unimpaired with SCD or MCI)

A total of 96 individuals of the β-AARC cohort were included in the present study, 88 cognitively unimpaired with SCD and 8 with MCI. Among them, 18 (18.75%) had a CSF profile of AD, while 78 (81.25%) did not (Table S3). Plasma UGOT p-tau217 was higher in Aβ+ versus Aβ-individuals (P<0.001, Fig. 4a) and had a discrimination accuracy between the two groups of 0.91 (Fig. 4b). Plasma UGOT p-tau217 was inversely correlated with CSF Aβ42/40 ratio (R=-0.52, P<0.001; Fig 4c) and positively correlated with CSF p-tau181 (R=0.37, p<0.001; Fig. 4d).

**Figure 4.**
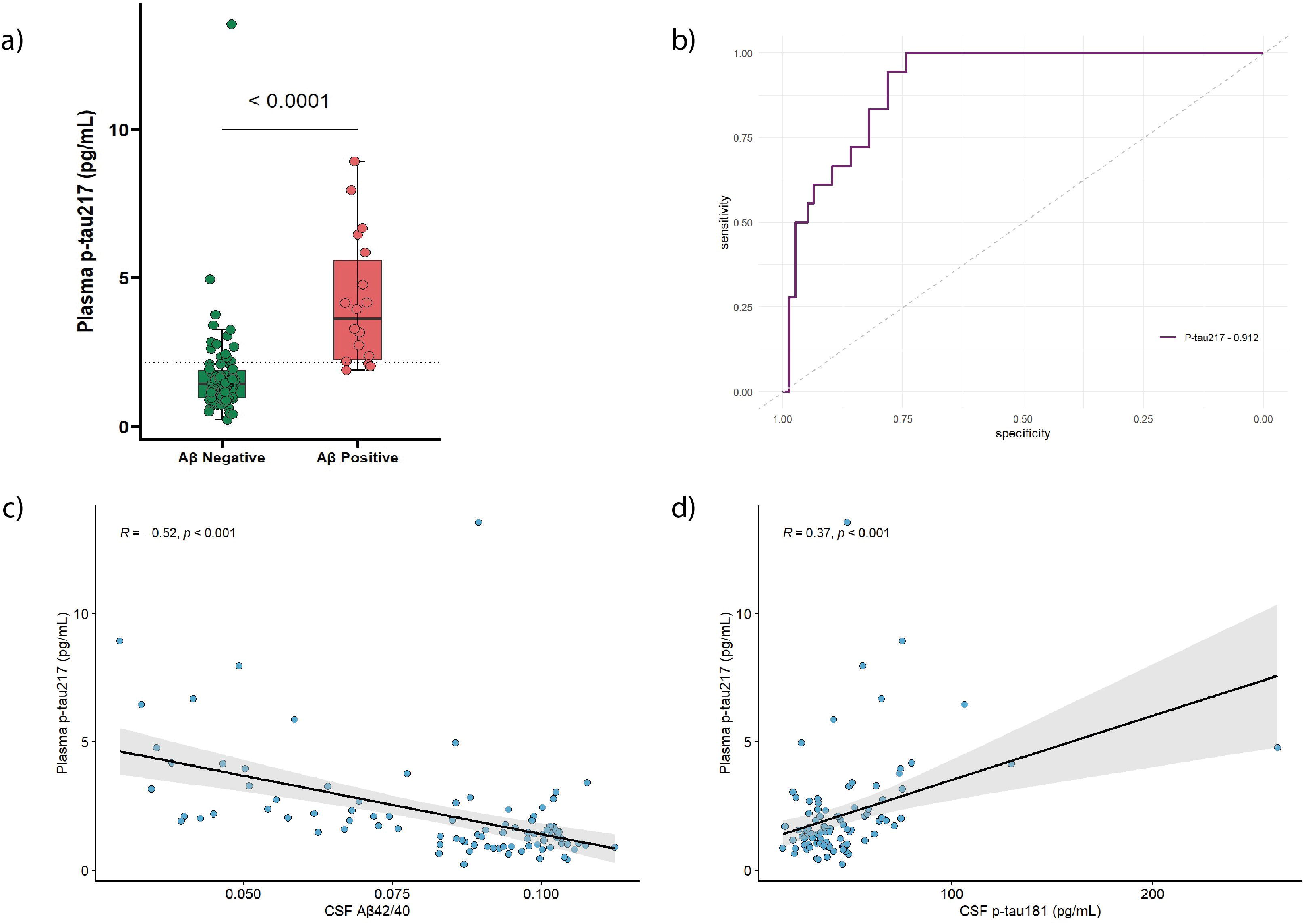
Plasma UGOT p-tau217 identifies amyloid positivity in CSF in individuals experiencing SCD. *(4a-b)* In the B-AARC cohort, plasma UGOT p-tau217 identified CSF Aβ+ in individuals experiencing SCD with high accuracy (AUC: 0.91). *(4c-d)* Plasma UGOT p-tau217 negatively correlate with the Aβ42/40 ratio in CSF (R=-0.52, p<0.001) and positively correlated with p-tau181 in CSF (R=0.37, p<0.001).

### 3.6 Comparison of UGOT p-tau217 with plasma p-tau217 by IP-MS

We further evaluated the performance of UGOT p-tau217 by comparing it with an immunoprecipitation-mass spectrometry (IP-MS) method for the measurement of p-tau217 in plasma^32^. While the UGOT p-tau217 assay measures p-tau217 in regular plasma samples, the IP-MS method is based on immunoprecipitation using three phosphorylation-independent tau antibodies to enrich for tau variants, followed by trypsination so that the peptide tau212-221 phosphorylated at position 217 can be measured. Despite these methodological differences, we observed a very strong correlation between the two assays (R=0.87, p<0.001, Fig 5).

**Figure 5.**
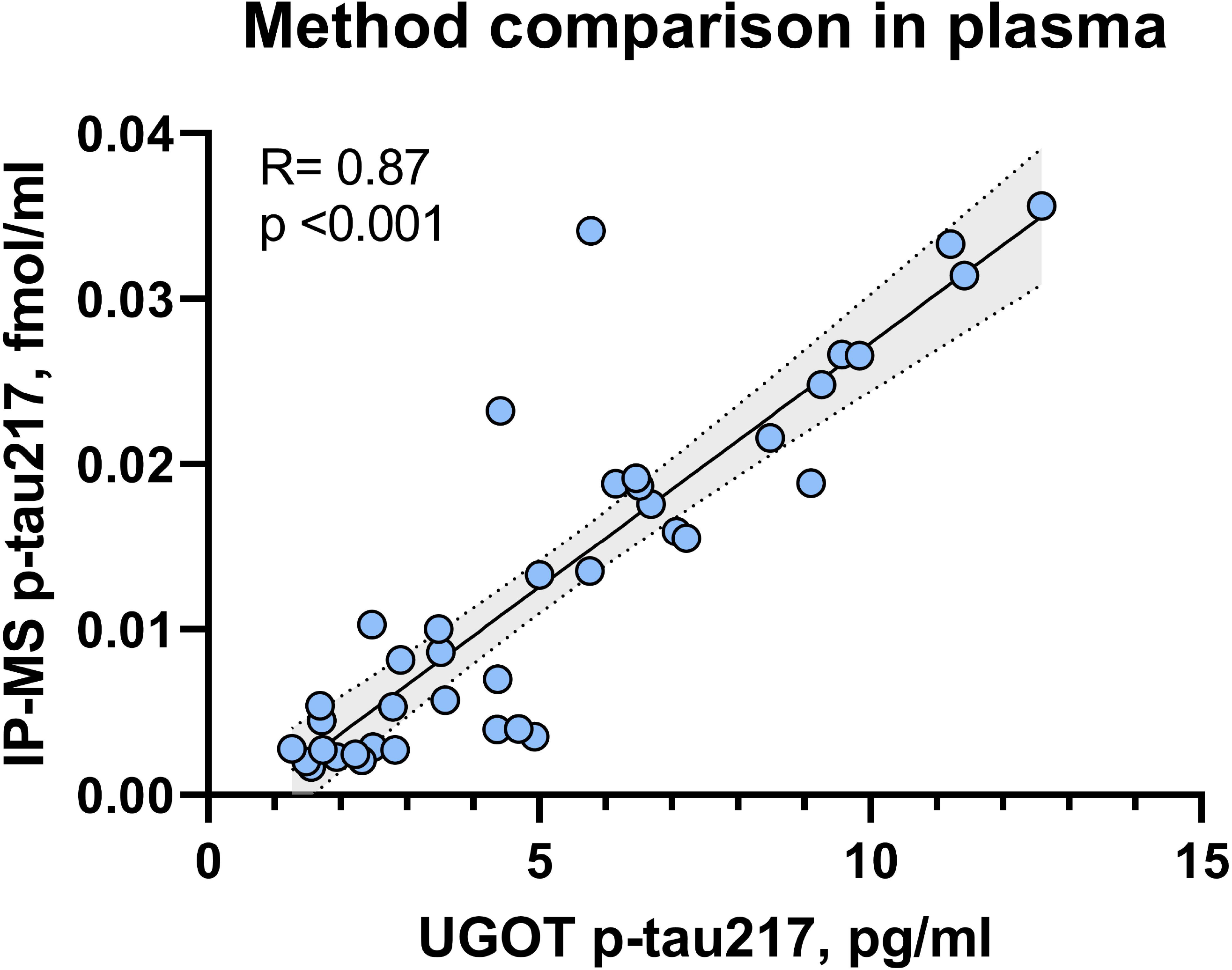
Plasma UGOT p-tau217 strongly correlates with p-tau217 measured by IP-MS. Comparison UGOT p-tau217 with p-tau217 measured by IP-MS in plasma. UGOT p-tau217 assay was strongly correlated with another method that uses IP-MS technology to quantify p-tau levels in plasma (R=0.87, p<0.001).

## 4. Discussion

Traditional clinical measures often have limited sensitivity to detect subtle changes, particularly in the early stages of the AD continuum. Blood-based tests could enable early diagnosis, facilitating timely and targeted therapeutic interventions to delay or prevent the onset of cognitive decline. Additionally, the accessibility and affordability of blood-based testing make them a practical tool for widespread screening and monitoring, potentially useful in AD clinical trials and drug development.

In this study, we have described a new ultrasensitive immunoassay for plasma p-tau217. We selected cohorts with different designs and target populations, allowing us to obtain information from various potential contexts of use. Some focused on early changes in cognition at the population level (MYHAT), by Aβ PET positivity (Pittsburgh) or CSF Aβ42/40 positivity (Discovery and B-AARC). The clinical performance, which focused on cohorts of individuals with emerging evidence of Aβ pathology, showed good associations with Aβ pathology assessed with either Aβ PET uptake or CSF Aβ42/40 ratio across four independent cohorts. In addition, the UGOT p-tau217 assay outperformed other plasma biomarkers including p-tau181, p-tau231, and GFAP, indicating that plasma p-tau217 might be a superior predictor of Aβ pathology in agreement with previous reports^15, 17^. Moreover, the UGOT p-tau217 assay was strongly correlated with another method that uses IP-MS technology to quantify p-tau217 levels in plasma. This suggests that our immunoassay-based method which has the advantage of measuring p-tau217 in regular plasma samples has equivalent performance as the method that first enriches for the target analyte through immunoprecipitation. Together, the novel UGOT p-tau217 method has a very high potential as a biomarker for detecting Aβ pathology, even in the early stages of the disease continuum.

Whilst previous studies have shown good biomarker accuracies for plasma p-tau217^17, 20^, performances among CU participants in population-based cohorts have been scanty and in the few available papers the results have been less consistent ^6, 12, 34^. This can be mainly attributable to the weak analytical performance (e.g., lack of quantifiable signals, poor precision) of first-generation p-tau217 assays in individuals with very low concentrations including CU participants and those in early symptomatic stages^2, 3^. However, detection of incipient Aβ pathology in the early stages of the AD continuum is crucial not only for early disease identification but also to determine eligibility for the recently approved anti-amyloid therapies. The high performances of the UGOT p-tau217 corroborate the results from previous studies^15, 17, 19^. The diagnostic performance of UGOT p-tau217 in early AD makes it a suitable candidate as first-in-line diagnostic test for patients with suspected AD pathology, regardless of the cognitive status.

Even though plasma p-tau217 assays on different platforms from some biotechnology/pharmaceutical companies are expected to become commercially available in the next few months, having an assay developed completely from an academic source removes restrictions such as the need to have the results cleared by the company in question before data can be published. Moreover, the companies tend to select cohorts with clinical and biomarker characterization that align with their commercial interests, limiting access to these assays. Getting rid of these limitations by providing access to a plasma p-tau217 assay with pure academic interests opens the possibility for assay testing an optimization in real-life scenarios. The UGOT p-tau217 assay presents an opportunity to fill the gap between academic research and clinical applications by ensuring that the new assay is available to academic partners for research and easily accessible for clinical settings.

Plasma UGOT p-tau217 had a stronger correlation with BD-tau than NfL when considering neurodegeneration biomarkers, which can be explained by the AD specificity of BD-tau over NfL^31^. Despite all three biomarkers showing alterations in AD, plasma p-tau217 and BD-tau reflect related biological processes that are more specific to AD compared with NfL.

In conclusion, we have the described the analytical and clinical performance of the novel plasma UGOT p-tau217 assay, the first one available from an academic centre. Given its high performances to identify individuals with abnormal Aβ PET scans and strong associations with cognitive performance and other CSF/blood biomarkers demonstrated across four independent cohorts, the method will be critical to screen for Aβ pathology in populations with or without cognitive impairment. As new anti-Aβ therapies become available for AD, well-validated blood biomarkers will be needed to screen patients for treatment eligibility and monitoring. Furthermore, since the field is moving toward earlier disease detection, blood biomarkers are critical for population screening for the identification and evaluation of both cognitively impaired and unimpaired older adults who may have biological evidence of Aβ pathology. The UGOT p-tau217 will be crucial to addressing these needs.

## Supporting information

https://www.editorialmanager.com/adj/download.aspx?id=394097&guid=30a5185a-ff81-4a14-9ce6-072af6e6a9ff&scheme=1

## Data Availability

All data produced in the present study are available upon reasonable request to the authors

## Acknowledgements

This publication is part of the BBRC’s β-AARC study, the MYHAT study and the Pittsburgh study. The authors would like to express their most sincere gratitude to the project participants, without whom this research would have not been possible. Authors would like to thank Altoida for kindly supporting the CSF AD core biomarker characterisation of β-AARC study participants.

## Consent Statement

The Discovery cohort used de-identified leftover clinical samples was approved by the ethics committees at the University of Gothenburg (#EPN140811).The Pittsburgh and MYHAT Cohorts were approved by the University of Pittsburgh Institutional Review Board and all participants gave written informed consent. The β -AARC study was approved by the independent ethics committee ‘Parc de Salut Mar’, Barcelona, and registered as Clinicaltrials.gov (identifier: NCT04935372). All participants gave written informed consent.

## Strengths and Limitations

Strengths of this study include the development and analytical validation of the plasma UGOT p-tau217 assay, and the multicentric evaluation of its associations with CSF AL42/40, Aβ PET and other blood-based biomarkers in independent cohorts across the AD continuum. Limitations include the lack of longitudinal UGOT p-tau217 data and neuropathology information.

