## Supplementary material for "A novel ultrasensitive assay for plasma p-tau217: performance in individuals with subjective cognitive decline and early Alzheimer’s disease": https://www.editorialmanager.com/adj/download.aspx?id=394097&guid=30a5185a-ff81-4a14-9ce6-072af6e6a9ff&scheme=1

**Supplementary Table 1.** Demographic table for the Discovery cohort

|  | **Discovery cohort** | |
| --- | --- | --- |
|  | **Control** | **AD** |
| **Sample size** | 20 | 20 |
| **Age, y** | 59.1 ±13.0* | 69.1 ±5.4 |
| **Sex, F, *n (%)*** | 12 (60%) | 14(70%) |
| **CSF Aβ42, pg/ml** | 902 ±231.9* | 397.3 ±106.3 |
| **CSF t-tau (Innotest), pg/ml** | 243.3 ±74.4* | 675.3 ±102.7 |
| **CSF p-tau181 (Innotest), pg/ml** | 36.4 ±11.73* | 86.3 ±27 |
| **CSF p-tau217, pg/ml** | 26.8 ±10.1* | 189.6 ±107 |
| **Plasma p-tau217, pg/ml** | 1.2 ±0.7* | 7.4 ±3.6 |

*Significantly different compared with the AD group (P<0.0001). Mann Whitney test.

**Supplementary table 2.-** Demographic characteristics of the MYHAT cohort

|  | **0 (N=40)** |  | **0.5 (N=39)** |  |  |  | **Overall (N=79)** | **p-value** |
| --- | --- | --- | --- | --- | --- | --- | --- | --- |
| **Age, y** |  |  |  |  |  |  |  |  |
| Mean (SD) | 80.2 (5.81) |  | 85.8 (4.72) |  |  |  | 83.0 (5.97) | < 0.001 |
| **Sex** |  |  |  |  |  |  |  |  |
| Female (%) | 30 (75.0) |  | 30 (76.9) |  |  |  | 60 (75.9) | 1 |
| **Education, y** |  |  |  |  |  |  |  |  |
| Mean (SD) | 13.7 (2.46) |  | 12.7 (2.71) |  |  |  | 13.2 (2.62) | 0.09 |
| **MMSE** |  |  |  |  |  |  |  |  |
| Mean (SD) | 28.3 (1.61) |  | 24.3 (5.75) |  |  |  | 26.3 (4.62) | < 0.001 |
| **Plasma p-tau217 (pg/mL)** |  |  |  |  |  |  |  |  |
| Mean (SD) | 2.56 (2.30) |  | 4.90 (3.37) |  |  |  | 3.71 (3.09) | < 0.001 |
| **Plasma p-tau181 (pg/mL)** |  |  |  |  |  |  |  |  |
| Mean (SD) | 2.32 (1.40) |  | 3.03 (1.54) |  |  |  | 2.67 (1.51) | 0.03 |
| **Plasma GFAP (pg/mL)** |  |  |  |  |  |  |  |  |
| Mean (SD) | 163 (71.7) |  | 259 (171) |  |  |  | 211 (138) | < 0.001 |
| **Plasma BD-tau (pg/mL)** |  |  |  |  |  |  |  |  |
| Mean (SD) | 13.5 (9.27) |  | 18.9 (14.3) |  |  |  | 16.2 (12.3) | 0.05 |

**Supplementary table 3.-** Demographic characteristics of the Pittsburg cohort

|  | | **Aβ Negative (N=72)** | | **Aβ Positive (N=21)** | | **Overall (N=93)** | | **p-value** |
| --- | --- | --- | --- | --- | --- | --- | --- | --- |
| **Age, y** | |  | |  | |  | |  |
| Mean (SD) | | 63.2 (8.76) | | 72.9 (6.72) | | 65.4 (9.25) | | < 0.001 |
| **Sex** | |  | |  | |  | |  |
| Female (%) | | 46 (63.9) | | 9 (42.9) | | 55 (59.1) | | 0.141 |
| MMSE/MoCA, | |  | |  | |  | |  |
| Mean (SD) | | 28.5(1.31)/23.98(3.68) | | 27.75(2.65)/25.86(2.48) | | 28.30(1.79)/24.38(3.49) | | 0.313/0.08 |
| **Plasma p-tau217 (z-score)** | |  | |  | |  | |  |
| Mean (SD) | | -0.282 (0.531) | | 1.15 (1.50) | | 0.0419 (1.03) | | < 0.001 |
| **Plasma p-tau181 (z-score)** | |  | |  | |  | |  |
| Mean (SD) | | -0.0744 (0.999) | | 0.326 (1.04) | | 0.0160 (1.02) | | 0.126 |
| **Plasma p-tau231 (z-score)** |  | |  | |  | |  | |
| Mean (SD) | | -0.0290 (1.05) | | 0.122 (0.908) | | 0.00508 (1.02) | | 0.522 |

**Supplementary Table 4.** Demographic characteristics of the β-AARC cohort

|  | **non-AD CSF profile**  **(n = 78, 81.25%)** | **AD CSF profile**  **(n = 18, 18.75%)** | **Total**  **(n = 96)** | ***P* value** | ***η_p_^2^*** |
| --- | --- | --- | --- | --- | --- |
| **Age,** years | 65.9 (5.50) | 72.6 (5.20) | 67.1 (6.03) | <0.001 | - |
| **Female,** n (%) | 42 (53.8%) | 11 (61.1%) | 53 (55.2%) | 0.767 | - |
| **Education,** years | 15.0 [12.0-18.0] | 12.5 [10.5-15.8] | 15.0 [12.0-18.0] | 0.025 | - |
| **MMSE** | 29.0 [27.2-29.0] | 28.0 [27.0-29.8] | 29.0 [27.0-29.0] | 0.580 | - |
| **AD CSF core biomarkers** (Lumipulse) | | | | | |
| **Aβ42/40** | 0.10 [0.09-0.10] | 0.04 [0.04-0.05] | 0.09 [0.07-0.10] | <0.001 | 0.668 |
| **p-tau181** (pg/ml) | 34.7 [28.7-45.6] | 65.3 [59.5-75.4] | 37.8 [29.4-50.0] | <0.001 | 0.262 |
| **t-tau** (pg/ml) | 266 [220-342] | 476 [388-510] | 285 [228-378] | <0.001 | 0.177 |
| **Plasma biomarkers** (University of Gothenburg, in-house) | | | | | |
| **UGOT p-tau217** (pg/ml) | 1.43 [0.96-1.89] | 3.62 [2.23-5.58] | 1.56 [1.02-2.35] | <0.001 | 0.183 |

Data are expressed as Mean (*M*) and Standard Deviation (*SD*) [age], Median (M) and interquartile range (Q1-Q3) [MMSE, AD CSF core biomarkers and plasma p-tau217] or number of participants (n) and percentage (%) [sex]. AD CSF profile was defined by a CSF Ab42/40 ratio < 0.062 (as measured by Lumipulse G600II, Fujirebio). CSF measurements were not available for 2 participants out of the 96 (2.1%) of the Roche NTK.

*P* values tested the difference between AD CSF core biomarkers profile groups and were computed with a *t* test (age), a Mann-Whitney *U* test (MMSE, AD CSF core biomarkers and plasma p-tau217) or a Chi square (sex).

Most participants were SCD, but 8 of them were MCI, with 5 classified as A- and 3 as A+ according to the aforementioned cut-off. This segregation was not statistically significant at 0.05 (p = 0.1682) after performing a Fisher exact test.

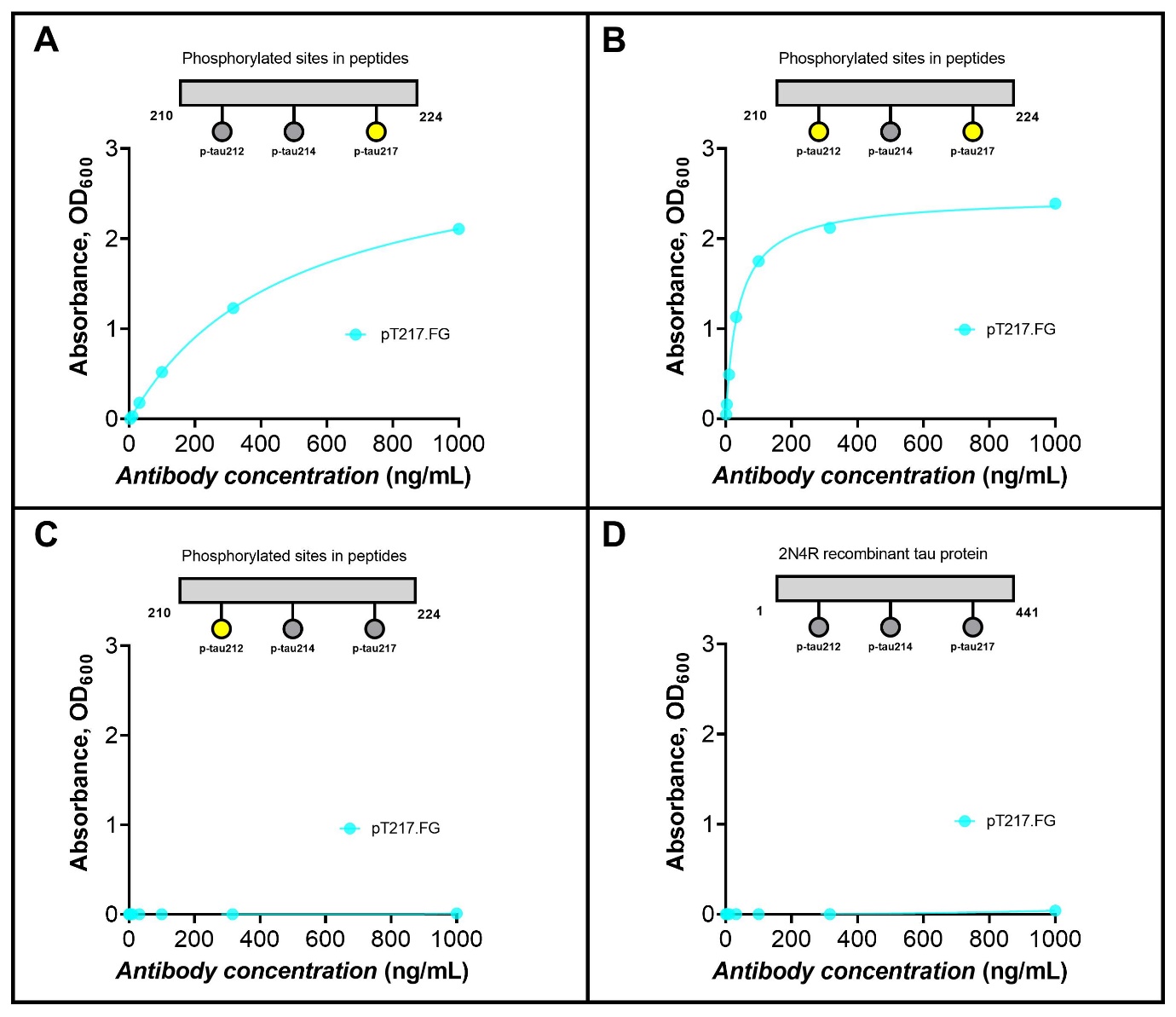

**Supplementary Fig. 1. Antibody validation.** A sheep monoclonal antibody was generated selectively against the p-tau217 epitope and avoids binding to any other phospho epitope in the region 210-220. The results of the direct ELISA show that our pT217 antibody online binds in presence of phosphorylation of threonine 217. In absence of the p-tau217 epitope the antibody shows no binding even in presence of 212 phosphorylation.

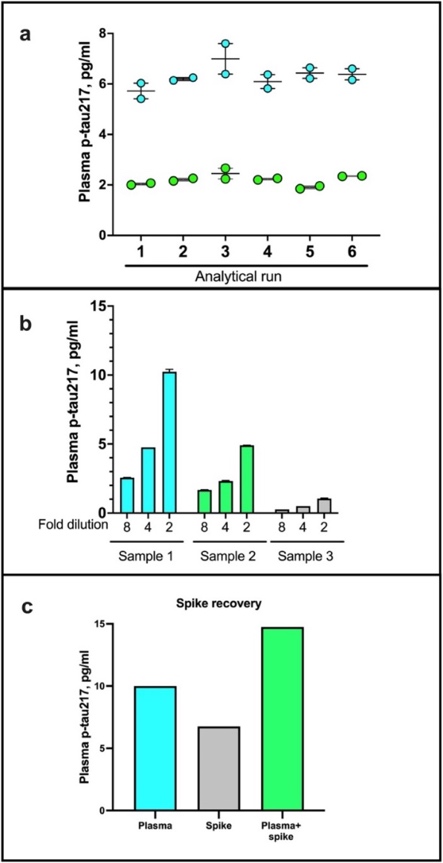

**Supplementary Fig. 2. Technical validation of the novel assay to measure p-tau217 in blood.** (a)*Within- and between-run stability*. The concentrations of two separate plasma samples were measured in duplicates in up to six independent analytical runs are shown, to depict day-to-day stability of the p-tau217 assay. **(b)** *Dilution linearity*. The panel shows serial dilutions of three different plasma samples with the assay diluent. Compared with sample aliquots diluted two-fold, those diluted four-fold gave approximately 50% less signal for p-tau217. The trend was the same when comparing four- and eight-fold diluted samples. **(c)** *Spike recovery.* samples diluted 1:2 as well as the assay diluent were each ‘spiked’ with *in vitro* phosphorylated recombinant full-length tau-441 (#TO8-50FN, SignalChem) and levels in each sample were measured with our p-tau217 assay. Figure 2c shows the concentrations for the unspiked plasma sample, the spike sample alone, and the plasma + spike sample together. The lower limit of quantification for the assay (LLOQ) was estimated to be 0.08 pg/mL. LLOQ was calculated by serially diluting the highest assay calibrator point (53.7 pg/mL) two-fold (and in duplicates) and setting the LLOQ as the calibrator point immediately preceding the first concentration where the coefficient of variation (CV) was 20% or above.
